# Prevalence of cancer susceptibility variants in patients with multiple Lynch syndrome related cancers

**DOI:** 10.1101/2021.02.22.21250719

**Authors:** Yoon Young Choi, Su-Jin Shin, Jae Eun Lee, Lisa Madlensky, Seung-Tae Lee, Ji Soo Park, Jeong-Hyeon Jo, Hyunki Kim, Daniela Nachmanson, Xiaojun Xu, Sung Hoon Noh, Jae-Ho Cheong, Olivier Harismendy

**Affiliations:** Department of Surgery, CHA University School of Medicine, Korea; Department of Surgery, Yonsei University Health System, Yonsei University College of Medicine, Seoul, Korea; Yonsei Biomedical Research Institute, Yonsei University Health System, Yonsei University College of Medicine, Seoul, Korea; Department of Pathology, Yonsei University Health System, Yonsei University College of Medicine, Seoul, Korea; Hereditary Cancer Clinic, Yonsei University Health System, Yonsei University College of Medicine, Seoul, Korea; Department of Laboratory Medicine, Yonsei University Health System, Yonsei University College of Medicine, Seoul, Korea; Department of Medicine, Yonsei University Health System, Yonsei University College of Medicine, Seoul, Korea; Moores Cancer Center, University of California San Diego School of medicine, San Diego, California, USA; Department of Family Medicine and Public Health, University of California San Diego School of medicine, San Diego, California, USA; Division of Biomedical Informatics, Department of Medicine, University of California San Diego School of medicine, San Diego, California, USA; Bioinformatics and Systems Biology Graduate Program - University of California San Diego School of medicine, San Diego, California, USA

**Keywords:** cancer susceptibility variant, multiple primary cancer, lynch syndrome, genomics, germline variant

## Abstract

**Background:** Along with early onset cancers, multiple primary cancers (MPCs) are likely resulting from increased genetic susceptibility; however, the associated predisposition genes or prevalence of the pathogenic variants genes in MPC patients are often unknown.

**Methods:** We screened 71 patients with MPC of the stomach, colorectal, and endometrium, sequencing 65 cancer predisposition genes. A subset of 19 patients with early onset MPC of stomach and colorectum were further evaluated for at DNA repair and cancer related genes using both normal (germline) and tumor (somatic) whole exome sequencing.

**Results:** Among 71 patients with MPCs, variants predicted to be pathogenic were observed in 15 (21.1%) patients and affected Lynch Syndrome (LS) genes: *MLH1* (n=10), *MSH6* (n=2), *PMS2* (n=2), and *MSH2* (n=1). All carriers had tumors with high microsatellite instability and 13 of them (86.7%) were early-onset, consistent with LS. In 19 patients with early-onset MPCs, loss of function (LoF) variants in *RECQL5*, including a rare East-Asian specific variant, were more prevalent in non-LS MPC than in matched sporadic cancer patients (OR=31.6, p=0.001). Additional evaluation of bi-allelic alterations in the tumor correctly identified LS genes in LS patients and candidates genes in non-LS patients including high-confidence LoF variants in 2 patients, *FANCG* (c.307+1G>C) and *CASP8* (p.R221Sfs*17) both accompanied by somatic loss of heterozygosity in a gastric and a colorectal tumor, respectively.

**Conclusions:** The results suggest that genetic screening should be considered for synchronous cancers and metachronous MPCs of the LS tumor spectrum, particularly in early-onset patients. Susceptibility variants in non-LS genes for MPC patients may exist, but evidence for their role is more elusive than for LS patients.

## Background

The identification of inherited DNA variants in cancer predisposition genes (CPG) is clinically important for both cancer prevention and treatment as it can reduce cancer morbidity and mortality in individuals carrying loss of function (LoF) variants. [1] The prevalence of LoF variants in CPG is lower than 1% of the healthy population and 5-8% in patients diagnosed with cancer. [2-4] LoF variants in CPG are therefore rare. Furthermore, surveillance screening has risks such as overdiagnosis, anxiety due associated with the risk of false positive, or limited effectiveness due to uncertain variants annotation and significance. Thus, continued research supporting more effective and precise genetic screening guidelines is needed. [1]

Multiple primary cancers (MPCs), referring to two or more histologically distinct cancers diagnosed in one individual, is gradually recognized as an important medical problem with a reported frequency of 2-17% in the cancer population. [5, 6] Along with early-onset cancer, MPC have been regarded as high-risk of heritability [7] and those patients therefore represent excellent candidates for genetic screening. However, limited data is available for the prevalence and type of LoF variants in the CPG of these patients.

Gastric cancer (GC) and colorectal cancer (CRC) are the two most common cancers in Korea, with 5-year survival up to 70% [8] and their combination is the most common instance of MPCs, representing ∼2% of Korean GC or CRC cases. [9, 10] About 15% of each of the cancer types can be characterized by elevated microsatellite instability (MSI) caused by mismatch repair (MMR) deficiency. [11-15] Together with Endometrial cancer (EC) - another cancer characterized by MSI [16] - GC and CRC are the main cancer types associated with Lynch Syndrome (LS), caused by LoF variants in MMR genes, resulting in up to an 80% lifetime risk of cancers. [17, 18] In particular, 2-5% of CRC and EC, as well as ∼15% of early-onset CRC are thought to be associated with LS. [19, 20] MPCs with cancer types in the spectrum of LS therefore represent some of the strongest candidates for genetic screening, as the prevalence of LS among MPC is unknown.

Here we present the results of the genetic screening of 71 patients with MPC of the stomach, colorectal and endometrium. We evaluate the distribution and prevalence of LoF variants in 65 CPGs, including LS susceptibility genes, as a function of age and clinicopathological features. We further characterize inherited coding variants focusing on DNA repair and cancer related genes in 19 early-onset MPC (eoMPC) patients, proposing candidate MPC susceptibility genes based on the analysis of LoF variants, their relative prevalence and association with the mutational landscape of the associated tumors.

## Methods

### Population

We performed a retrospective population-based study targeting patients who were treated for two or more cancers in the stomach, colorectum, or endometrium at Severance Hospital, Yonsei University College of Medicine between January 2001 to December 2016. The patients were selected using following criteria: 1) two or more cancers were pathologically confirmed, 2) multiple tumors were histologically different, not a metastatic or recurrent tumor from one cancer, 3) normal tissues were available and histologically confirmed for sequencing. DNA was obtained from formalin fixed paraffin embedded (FFPE) normal and tumor tissues. Early-onset MPC (eoMPC) was defined by the diagnosis of a second cancer at age 55 or younger. The clinico-pathologic characteristics of the patients and tumors including age, sex, family history of cancer, MSI or MMR status of tumors were evaluated. For evaluating MSI status, two mononucleotide repeat markers (BAT25 and BAT26) and three dinucleotide repeat markers (D5S346, D2S123, and D17S250) were used by polymerase chain reaction, [21] and MSIsensor was used with cut-off of MSI score >3.5 in Whole Exome Sequencing (WES) data. [22] To evaluate protein loss of MMR gene, immunohistochemistry (IHC) was conducted in four MMR genes; *MLH1* (ready-to-use, clone M1, Roche, Indianapolis, IN, USA), *MSH2* (ready-to-use, clone G219-1129, Roche), *MSH6* (1:100, clone 44, Cell Marque, Rocklin, CA, USA), and *PMS2* (1:40, clone MRQ28, Cell Marque). An MMR-deficient (dMMR) tumor was defined as a tumor showing loss of expression of any of the four MMR proteins. If a tumor was classified as any one of MSI-high (MSI-H) or deficiency MMR (dMMR) it was considered as MSI-H. The concurrency of tumors was classified as synchronous tumors when the interval between tumors was less than 1 year, otherwise considered as metachronous tumors. This study was approved by Institutional Review Board (IRB) of Severance Hospital and University of California, San Diego (4-2017-0434, 191543)

### Germline multigene targeting panel

Germline DNA of patients with MPCs was evaluated using a customized targeted capture sequencing panel (OncoRisk, Celemics, Seoul, Korea) covering all coding sequences and intron-exon boundaries of the coding exons of known 65 CPG (Additional file 1, Table S1). [23] The germline variants were classified into pathogenic, likely pathogenic, variant of uncertain significance (VUS) and reported following the guidelines of the American College of Medical Genetics and Genomics 2017, [24] and pathogenic or likely pathogenic (P/LP) germline variant was regarded as CPG. Benign and likely benign variants were not considered in this study.

### Whole exome sequencing

#### Data generation

WES of normal and tumors was conducted for patients with eoMPCs in the stomach and colon. Genomic DNA was extracted from the confirmed normal and tumor tissues of FFPE. SureSelect sequencing libraries were prepared according to the manufacturer’s instructions (Agilent SureSelect All Exon V6 kit, Santa Clara, CA, USA), and HISEQ2500 sequencing system (Illumina^™^, San Diego, CA, USA) performed sequencing with read lengths of 2 × 100 bp. The statistics and quality metrics of normal and tumors were described in Additional file 1, Table S2 and S3.

#### Data analysis

The reads were aligned to hg19 reference genome by Burrows-Wheeler Aligner software (BWA 0.7.17) and duplicated reads were removed by Picard (MarkDuplicates) through bcbio-nextgen (v.1.2.3) [25] For germline and somatic analysis, variants were called by Genome Analysis Toolkit Haplotype joint caller (GATK v3.9) and mutect2, [26, 27] respectively. Variants were annotated by refGene using ANNOVAR, [28] CADD scores (CADD13 and CADDindel), [29] population allelic frequency in ExAC v3.0, [30] and membership to ClinVar. [31]

To discover novel CPG, we focused on high-confidence (TLOD>12, FS<10, SEQQ>60, MQε60, STRANDQ>40, DP>10), rare (minor allele fraction was < 0.01 in ExAC of Eastern Asian population), and damaging (exonic/splicing, non-synonymous variants with >20 of CADD score) variants in 382 genes of 11 cancer-relevant pathways, [32] and they were defined as LoF germline variants. High-confidence of LoF variants were predicted by LOFTEE plugin for Ensembl VEP (v.99). [33, 34] Copy number alterations of tumors compared to normal was estimated using CNVkit (V0.8) [35] The gene level copy number ratio was calculated as the weighted mean of all bins covered by the whole segment overlapping the gene. Genes had ε3 segments of copy number changes were included, and log2 copy number < −0.3 was defined as deletion. Somatic allelic imbalance (AI) in tumors of a given heterozygous germline variants were estimated using hapLOHseq. [36] Germline variants of tumors were obtained by GATK haplotype caller and filtering out variants were not observed in germline variants in normal sample. Phase of genotypes were estimated with a companion phasing utility with hapLOHseq. Event prevalence was set 0.1, and AI was defined when posterior probability of being in AI was over 90%. To assess loss of heterozygosity (LOH) of a given heterozygous germline variants in tumors as a secondary hit mechanism of inactivation of CPG, [32, 37] we evaluated germline and somatic bi-allelic alteration of corresponding tumors. When there was any one of somatic damaging mutation or AI event in candidate germline CPG, it was considered as bi-allelic alteration.

To compare the frequency of recurrent CPG in patients of eoMPCs to that of the cancer genome atlas (TCGA) cohort, germline data of East-Asian patients (≥80% of admixture) [38] with stomach cancer or colorectal cancer in TCGA cohort was used. [32, 39] Variants were filtered for rare and damaged variants similar to LoF germline variants in MPCs cohort. For recurrent variants and genes in eoMPCs cohort, the frequency was compared between eoMPCs cohort and selected TCGA cohort.

### Statistical analysis

Continuous variables were compared by two-sided Mann-Whitney test and categorical variables were compared by two-sided Fisher’s exact test. P<0.05 was considered as statistically significant and R version 3.6.1(R Foundation for Statistical Computing, Vienna, Austria) was used for generating figures and statistical analysis.

## Results

### Identification of Lynch Syndrome patients by multigene targeting panel

We investigated the prevalence of LoF variants in CPG using a cohort of 71 patients with MPCs. Fifty-four (76.1%) patients were diagnosed with GC and CRC (23 −42.6% synchronous) while 14 diagnosed with EC and CRC (N=13) or GC (N=1). Three patients had 3 or more types of cancers. Thirty (42.3%) patients were diagnosed with MPC before age 55 and classified as eoMPCs. A family history of cancer was more prevalent in eoMPCs: of the 56 patients for which family history was available, twenty-four (42.9%) patients had two or more first-degree relatives affected with any type of cancer, and 15 of them were eoMPCs (Odds Ratio [OR]=8.58, 2.21-39.59, *p*<0.001). Twenty-six (36.6%) patients had one or more MSI-H tumors, a higher proportion than the one observed in single-cancers of the same type (∼15%) [11-13, 16] (Supplemental File 1, Table S4). This suggests an important contribution of defects in MMR to MPC development. The germline DNA of the patients was sequenced using a multigene targeting panel of 65 CPGs. A total of 15/71 (21.1%) patients were found to carry variants predicted to be pathogenic or likely pathogenic (P/LP) and all the variants affected LS related genes (Table 1, Table S5 in Supplemental File 1): *MLH1* (n=10), *MSH6* (n=2), *PMS2* (n=2), and *MSH2* (n=1), a distribution comparable to the previous reports [17] One of recurrent P/LP variants of *MLH1* was c.1758dupC, a common variant in Korean LS patients. [40] There was a P/LP variant in *MSH2* (c.942+3A>T) that was reported as frequently *de novo* mutation [41] in a patient diagnosed with four types of cancer (GC, CRC, EC, and lung cancer). Of the remaining 56 of patients, 42 had VUS altering 43 genes, leaving the contribution of these genes to MPC undetermined.

**Table 1.** Summary of clinicopathologic characteristics of patients with multiple Lynch related primary cancers with pathogenic/likely-pathogenic germline variants by targeted panel.

| Case ID | Cancer type | Age at second diagnosis | Gene | Nucleotide change | Amino acid change | Mutation type | class† | MSI status |
| --- | --- | --- | --- | --- | --- | --- | --- | --- |
| dou_002 | CRC/GC | <55 | *MLH1* | c.1721T>C | p.Leu574Pro | missense | LP | MSI-H/MSI-H |
| dou_003 | GC/CRC | <55 | *MLH1* | c.1758dupC | p.Met587HisfsTer6 | frameshift_insertion | LP | MSI-H/MSI-H |
| dou_005 | CRC/GC | <55 | *MLH1* | c.208-1G>A | NA | splice_acceptor_variant | P | MSI-H/MSS |
| dou_006 | GC/CRC | <55 | *MLH1* | c.2041G>A | p.Ala681Thr | missense | LP | MSI-H/MSI-H |
|  |  |  | *BRCA1* | c.213-1G>A | NA | splice_acceptor_variant | P |  |
| dou_011 | GC/CRC | <55 | *MLH1* | c.790+2T>A | NA | splice_donor_variant | P | MSI-H/MSI-H |
| dou_016 | GC/CRC | <55 | *MLH1* | c.1758dupC | p.Met587HisfsTer6 | frameshift_insertion | LP | MSI-H/MSI-H |
| dou_017 | GC/CRC | <55 | *MLH1* | c.1559-2A>C | NA | splice | P | MSI-H/MSI-H |
| dou_047 | CRC/GC | >55 | *MSH6* | c.829G>T | p.Glu277Ter | splice_acceptor_variant | LP | MSI-H/NA |
| dou_056 | CRC/EC | >55 | *PMS2* | c.1738A>T | p.Lys580Ter | nonsense | P | MSI-H/MSS |
| dou_061 | CRC/EC | <55 | *MSH6* | c.3477C>G | p.Tyr1159Ter | nonsense | P | NA/MSI-H |
| dou_062 | CRC/EC | <55 | *PMS2* | c.943C>T | p.Arg315Ter | nonsense | P | MSI-H/MSI-H |
| dou_065 | CRC/EC | <55 | *MLH1* | c.67G>T | p.Glu23Ter | nonsense | P | MSI-H/MSI-H |
| dou_069 | CRC/EC/Klaskin | <55 | *MLH1* | c.67G>T | p.Glu23Ter | nonsense | P | MSI-H/MSI-H/MSI-H |
| dou_070 | EC/CRC/GC | <55 | *MLH1* | exon 7-9 deletion | NA | NA | LP | MSI-H/NA/MSI-H |
| dou_071 | CRC/GC/EC/Lung | <55 | *MSH2* | c.942+3A>T | NA | exon loss | LP | MSI-H/MSI-H/NA/MSI-H |

\* the order of MSI/MMR status is matched to cancer type
†: LP: likely pathogenic, P: Pathogenic

### Characteristics of LS patients

All 15 LS patients had at least one MSI-H tumor in contrast to 11/56 non-LS patients. LS patients were also more likely to have two or more first-degree relatives (9/10, OR=17.67, *p*=0.001, Additional file 1, Table S4 and S5). The association of the variants with tumor characteristics were further investigated. The loss of expression of the protein corresponding to the gene with P/LP variants was confirmed in all 19 MSI-H tumors investigated from all LS patients (Additional file 1, Table S6). The LS patients were more likely to be diagnosed with eoMPC (13/15, OR=14.31, p<0.001, Figure 1) but equally likely to present with synchronous tumors (6/15, OR=0.89, p>0.999). This suggests that consistent with Lynch syndrome, LS MPC patients are diagnosed earlier, but that the variants are unlikely to mediate the relative timing of the diagnosis. More importantly, these results showed that the majority of MPC patients do not have canonical LS mutations. Even restricting to 13 patients with eoMPCs and strong family history of cancer (two or more first-degree relatives over two successive generations), CPG were not altered in nearly half of the patients (6/13 Table S5 in the Additional file 1). The likely genetic cause of MPC in these patients remains undetermined.

**Figure 1.**
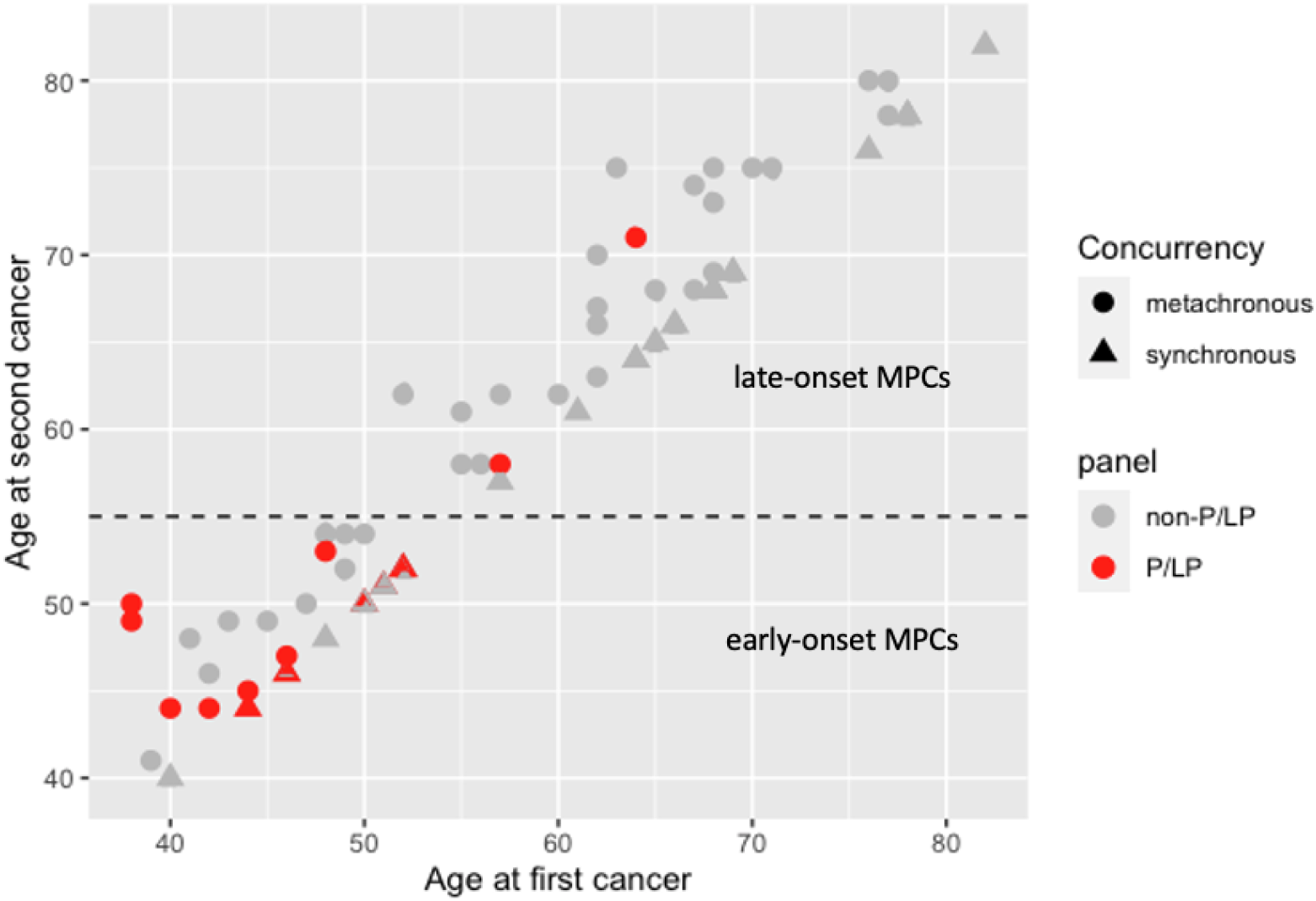
A scatter plot between age of first and second cancer by with/without pathogenic or likely pathogenic (P/LP) germline variants and concurrency of the tumors in each patient. There were 15 (21.1%) of patients with germline P/LP variants (red colors). Among early-onset multiple primary cancers (MPCs), 43.3% (13/30) patients had P/LP variants while only 4.9% (2/41) patients were related to P/LP variants in late-onset MPCs.

### Identification of candidate CPG in early-onset MPC

To identify additional variants and genes underlying a possible cancer susceptibility in non-LS patients, we sequenced the whole exome of 19 eoMPC patients with gastric and colorectal cancer which is the most common combination of MPCs in Korea. [9, 10] Seven of the patients were LS patients, as determined above, and used as controls for the CPG discovery process. We identified 111,842 high-confidence variants across all patients, of which 3,211 were rare (MAF<0.01) in East-Asian population and predicted to be damaging, affecting 2,675 genes. We investigated a set of 382 genes (referred to as cancer genes) involved in DNA repair, cancer susceptibility and progression (Additional file 1, Table S7). [32] Of those, 66 were altered by 82 variants, including 7 in the *MLH1* gene of the LS patients, therefore confirming the panel sequencing results and validating the initial WES variant calling and filtering approach.

For a given candidate CPG, we expect that the prevalence of LoF alterations observed in eoMPC cohort would be higher than in a matching sporadic cancer cohort. Despite some LS phenotypes being reported, [32] TCGA patients are mainly from sporadic cancers and we therefore selected 70 patents with gastric and colorectal and of East-Asian ancestry as a reference dataset. In these patients, we identified a total of 87 LoF variants affecting 47/382 cancer genes. We confirmed that LoF variants in *MLH1* were more prevalent in LS patients than in TCGA patients (7/7 vs 2/70 p<0.001 Table 2, Table S8 in the Additional file 1). Importantly, 5/7 of the *MLH1* LoF variants in LS patients were predicted to be high-confidence LoF variants, indicating that most, but not all, known pathogenic variants can be identified through this approach. *RECQL5* was the only gene significantly more altered in non-LS patients than in TCGA patients (4/12 vs 1/70, OR:31.66, *p*=0.0013, Table 2). In particular two of these patients had the same *RECQL5* variant (p.R441Q which is not seen in the TCGA patients. (*p*=0.02, Table S8 in the Additional file 1) The variant is rare, with a global minor allelic fraction of 3.6 × 10^−4^. Interestingly 89 of the 101 observed minor alleles (out of 278,974 total alleles observed) in the gnomAD dataset [42] belong to the East-Asian population, suggesting a possible Korean-specific effect.

**Table 2.** Genes affected by Loss of function variants in two or more patients with eoMPC at stomach and colon.

| Gene | N mutated cases |  |  |  | p-value*<br>(TCGA) | p-value+<br>(TCGA) |
| --- | --- | --- | --- | --- | --- | --- |
|  | non-Lynch<br>(n=12) | Lynch<br>(n=7) | total<br>(n=19) | TCGA_EAS<br>(n=70) |  |  |
| *MLH1* | 0 | 7 | 7 | 2 | >0.999 | **<0.0001** |
| *RECQL5* | 4 | 0 | 4 | **1** | **0.0013** | >0.999 |
| *EME2* | 2 | 0 | 2 | 1 | 0.0547 | >0.999 |
| *EP300* | 2 | 0 | 2 | 1 | 0.0547 | >0.999 |
| *MUTYH* | 2 | 0 | 2 | 1 | 0.0547 | >0.999 |
| *MSH3* | 2 | 0 | 2 | 2 | 0.1002 | >0.999 |
| *COL7A1* | 2 | 0 | 2 | 4 | 0.2108 | >0.999 |
| *PTCH1* | 2 | 0 | 2 | 7 | 0.6134 | >0.999 |

\* fisher's exact p-value for the frequency of each gene between non-lynch and TCGA
"+" fisher's exact p-value for the frequency of each gene between lynch and TCGA

### Bi-allelic alterations in corresponding tumors as a guide for CPG identification

According to Knudson’s two-hit model, [37] the wild-type allele of CPGs is frequently lost or mutated in the tumor. Such combination of inherited deleterious variants with somatic loss - referred to as bi-allelic alteration - can help prioritize candidate CPG. [32, 43] We sequenced the exome of 37 tumors from all 19 patients with eoMPC (Additional file 1, Table S3) and identified 35 somatic bi-allelic alterations by LoF mutations (N=5), loss of heterozygosity (N=27) or both (N=3) focusing on candidate germline LoF genes.

The *MLH1* gene was *somatically* altered in 13/15 tumors from LS patients and at least one tumor from each of the 7 LS patient. (Table 3). Importantly, 5/7 *MLH1* variants in LS patients were high-confidence LoF variants. This observation confirms the validity of the investigation of bi-allelic alteration to identify CPG, at least for high-confidence LoF variants in known predisposition genes. In contrast, the variants in the main candidate susceptibility gene in non-LS patients, *RECQL5*, were not high-confidence LoF variant and the genes was not affected in any of the 6 tumors from 4 variant carriers or any of the 22 tumors from all non-LS patients. Expanding the analysis to other candidate genes affected by LoF germline variants in non-LS patients, we identified 17 genes in 10 patients also affected by bi-allelic alterations in one or more tumors. Two were altered via mutations and 15 through LOH. Among these, 2 genes were affected by high-confidence LoF germline variants: *FANCG* (germline splice site variant, c.307+1G>C) in gastric tumor of dou_004 and *CASP8* (germline deletion variant, c.658_659del) in colorectal tumor of dou_010 (Table 3). Interestingly, this analysis also revealed somatic alterations of LS genes in absence of inherited LS mutation in a non-LS patient (dou_009) with both of MSI-H tumors in the stomach and colon. Indeed, somatic allelic imbalance was observed in *MLH1* (stomach and colon) and *PMS2* (stomach only) and the expression of both encoded proteins was lost from both tumors. Loss of one allele in the tumor would likely not be sufficient to impair MMR, raising the possibility of undetected germline LoF variants in these LS genes in this patient, perhaps through alterations not easily detected by WES. [44]

**Table 3.**
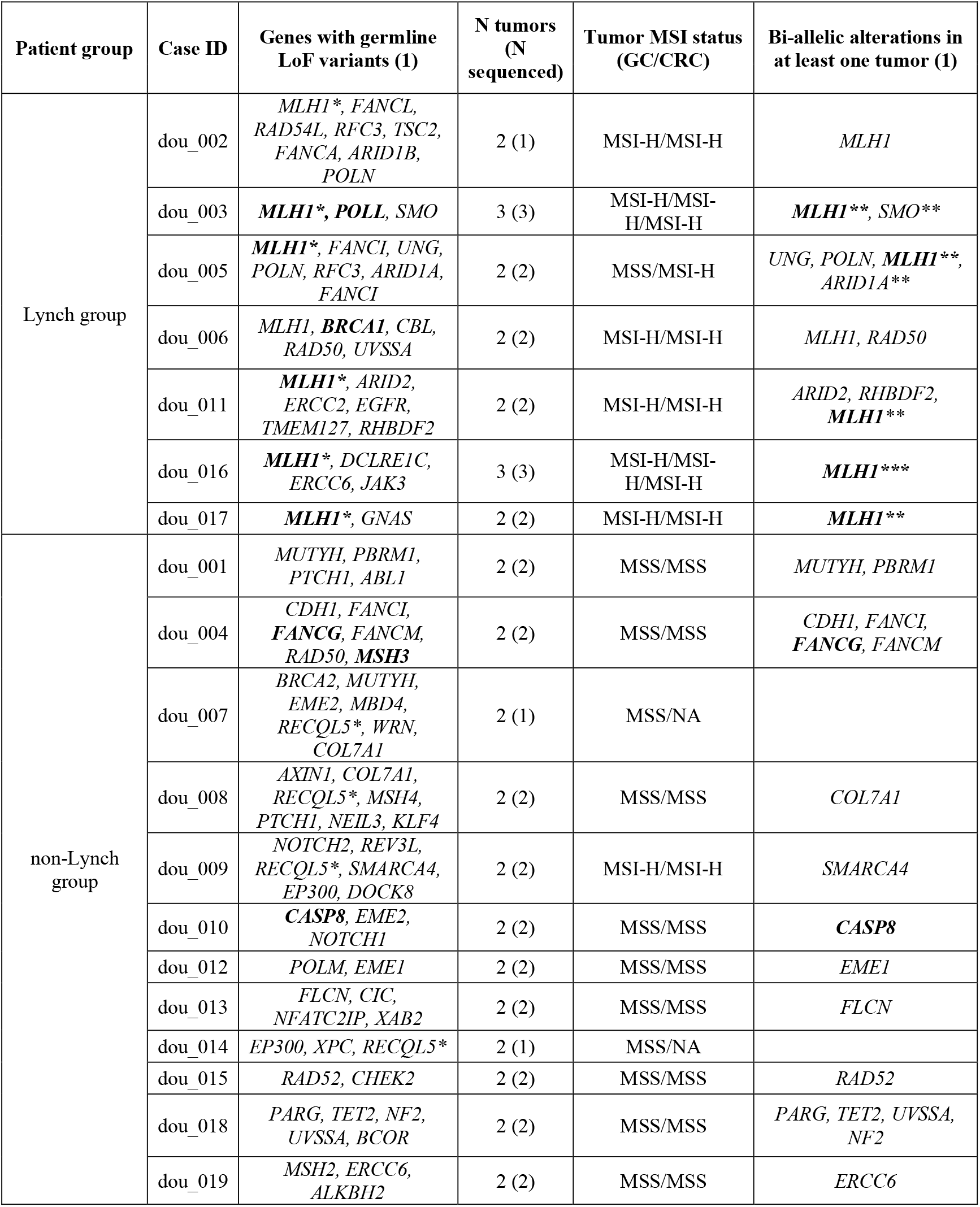
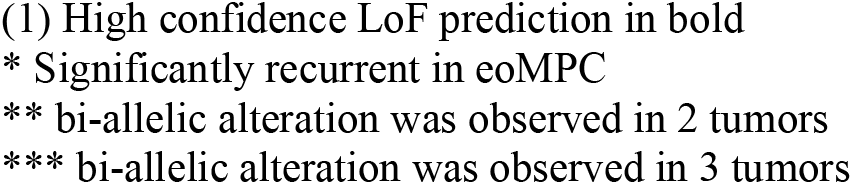
Summary of candidate loss-of-function germline candidate genes and the status of loss of heterozygocity of corresponding tumors.

| Patient group | Case ID | Genes with germline LoF variants (1) | N tumors (N sequenced) | Tumor MSI status (GC/CRC) | Bi-allelic alterations in at least one tumor (1) |
| --- | --- | --- | --- | --- | --- |
| Lynch group | dou_002 | *MLH1\**, *FANCL*, *RAD54L*, *RFC3*, *TSC2*, *FANCA*, *ARID1B*, *POLN* | 2 (1) | MSI-H/MSI-H | *MLH1* |
|  | dou_003 | *MLH1\**, *POLL*, *SMO* | 3 (3) | MSI-H/MSI-H/MSI-H | *MLH1\*\**, *SMO\*\** |
|  | dou_005 | *MLH1\**, *FANCI*, *UNG*, *POLN*, *RFC3*, *ARID1A*, *FANCI* | 2 (2) | MSS/MSI-H | *UNG*, *POLN*, *MLH1\*\**, *ARID1A\*\** |
|  | dou_006 | *MLH1*, *BRCA1*, *CBL*, *RAD50*, *UVSSA* | 2 (2) | MSI-H/MSI-H | *MLH1*, *RAD50* |
|  | dou_011 | *MLH1\**, *ARID2*, *ERCC2*, *EGFR*, *TMEM127*, *RHBDF2* | 2 (2) | MSI-H/MSI-H | *ARID2*, *RHBDF2*, *MLH1\*\** |
|  | dou_016 | *MLH1\**, *DCLRE1C*, *ERCC6*, *JAK3* | 3 (3) | MSI-H/MSI-H/MSI-H | *MLH1\*\*\** |
|  | dou_017 | *MLH1\**, *GNAS* | 2 (2) | MSI-H/MSI-H | *MLH1\*\** |
| non-Lynch group | dou_001 | *MUTYH*, *PBRM1*, *PTCH1*, *ABL1* | 2 (2) | MSS/MSS | *MUTYH*, *PBRM1* |
|  | dou_004 | *CDH1*, *FANCI*, *FANCG*, *FANCM*, *RAD50*, *MSH3* | 2 (2) | MSS/MSS | *CDH1*, *FANCI*, *FANCG*, *FANCM* |
|  | dou_007 | *BRCA2*, *MUTYH*, *EME2*, *MBD4*, *RECQL5\**, *WRN*, *COL7A1* | 2 (1) | MSS/NA |  |
|  | dou_008 | *AXIN1*, *COL7A1*, *RECQL5\**, *MSH4*, *PTCH1*, *NEIL3*, *KLF4* | 2 (2) | MSS/MSS | *COL7A1* |
|  | dou_009 | *NOTCH2*, *REV3L*, *RECQL5\**, *SMARCA4*, *EP300*, *DOCK8* | 2 (2) | MSI-H/MSI-H | *SMARCA4* |
|  | dou_010 | *CASP8*, *EME2*, *NOTCH1* | 2 (2) | MSS/MSS | *CASP8* |
|  | dou_012 | *POLM*, *EME1* | 2 (2) | MSS/MSS | *EME1* |
|  | dou_013 | *FLCN*, *CIC*, *NFATC2IP*, *XAB2* | 2 (2) | MSS/MSS | *FLCN* |
|  | dou_014 | *EP300*, *XPC*, *RECQL5\** | 2 (1) | MSS/NA |  |
|  | dou_015 | *RAD52*, *CHEK2* | 2 (2) | MSS/MSS | *RAD52* |
|  | dou_018 | *PARG*, *TET2*, *NF2*, *UVSSA*, *BCOR* | 2 (2) | MSS/MSS | *PARG*, *TET2*, *UVSSA*, *NF2* |
|  | dou_019 | *MSH2*, *ERCC6*, *ALKBH2* | 2 (2) | MSS/MSS | *ERCC6* |

- (1) High confidence LoF prediction in bold - \* Significantly recurrent in eoMPC - \*\* bi-allelic alteration was observed in 2 tumors - \*\*\* bi-allelic alteration was observed in 3 tumors

## Discussion

As expected, the present result confirmed that individuals with MPC, particularly early onset, have a higher likelihood of LS: MMR related P/LP germline variants were observed in 21% of MPC (15/71) and in 43% of eoMPC (13/30) patients, a proportion comparable to ∼15% of early-onset CRC. [20] The elevated extracolonic cancer risk following colorectal cancer in Lynch syndrome was reported (∼5 and 40 times of gastric and endometrial cancer risk compared to general population, respectively), [45] and this study supports those results. Therefore, due to the high prevalence of germline mutations individuals with MPC in the LS tumor spectrum should undergo germline testing, including when MSI status is unknown.

Because MSI-H was considered as a hallmarks of LS, universal screening of all CRC and EC has been recommended. [46, 47] In addition, MSI screening for gastric cancers in regions where gastric cancer is highly prevalent like Korea is a way to identify individuals and families who can benefit from germline testing of LS genes, so that surveillance and risk reducing interventions can be undertaken along with cascade testing of family members. At the same time, MSI testing is valuable for other types of solid cancers for other purpose: MSI-H is a biomarker for response to immune checkpoint inhibitors that is a breakthrough for treating advanced solid cancers regardless of its origin. [48] As MSI-H is predictive of LS across a broad tumor spectrum, [49] screening of tumor MSI status in all patients with an initial cancer, especially for LS spectrum tumors and early-onset cancer, and consecutive germline testing for patients with MSI-H tumor will help refine the diagnosis, giving an opportunity to diagnose LS and avoid or detect the second cancer as early as possible.

Genetic analysis of families with high occurrence of cancer using CPG panels or exome sequencing has become standard to identify the underlying cancer susceptibility variants. But penetrance, interaction with environmental factors, and size of the pedigree may affect power of such linkage studies. [1, 50-52] Evaluating both germline and somatic alterations is another reasonable approach to find CPG. [32, 43] The present results showed that characterizing recurrent LoF genes, truncating variants and loss of heterozygosity are useful to prioritize candidate CPG; any combination of them unambiguously identified germline *MLH1*, a known CPG in LS patients, in multiple patients clinically diagnosed with LS. It suggests that MPC is a strong phenotype to find CPG, therefore this approach would be worthy to be expanded to other combinations of MPCs. However, the types of MPCs would be different by geographical regions or ancestry: region-specific environmental factors can contribute to the oncogenesis and lead to different cancer types within the LS spectrum. For instance, MPCs including GC could be a hallmark of LS in Korea but not in the United States where GC incidence is much lower. This could explain why GC risk in LS may have been underestimated. [17, 53] Therefore, geographical and/or ancestry specific cancer epidemiology needs to be accounted for in the genetic screening guidelines.

In non-LS patients with eoMPC, however, evidence of germline susceptibility was more elusive. *RECQL5* was a frequently recurrent gene and variant (p.R441Q) compared to East Asian sporadic cancer population (TCGA), however there was no evidence of somatic LOH in the corresponding tumors. *RECQL5* variants have been associated with susceptibility to breast cancer, [54] and were observed in *CDH1* negative hereditary diffuse gastric cancer family [55] ; and its expression in GC is low, [56] and its role in MPC susceptibility is worthy of further investigations. *CASP8* and *FANCG* were high-confidence LoF germline gene with bi-allelic alteration in CRC and GC, respectively. *CASP8* encodes a member of caspase family and play a role in apoptosis, and some of its polymorphism have been reported as susceptibility to various cancers including CRC. [57-59] *FANCG* is a gene encodes Fanconi anemia (FA) group G protein, and related to FA DNA damage repair pathway that a possible germline predisposition to some sporadic cancers. [60-62] Thus, the *CASP8* and *FANCG* variants identified in these two MPC cases could underlie their disease susceptibility though additional functional studies would be necessary to demonstrate their pathogenicity. [63-66]

Despite evaluating all inherited coding variants in cancer genes and somatic changes in the corresponding tumors, we did not find a clear inherited predisposition that caused multiple cancers in over half of patients with eoMPC, even with strong family cancer history. Not only CPG, but also environmental factors such as smoking and alcohol also increase the risk of cancers. [67-69]

Our study has inherent limitations. The present results did not cover epigenetic changes affecting candidate CPG in tumors, and methylation of the *MLH1* promoter in particular is known to be a mechanism for somatic loss of function in ∼20% of CRC. [70] Furthermore, we restricted the analysis to 382 well studied cancer genes, more likely to impact cancer susceptibility, therefore leaving the possibility to miss un-expected CPG. The complete analysis of whole exomes to identify rare disease susceptibility variants would indeed be intractable for the cohort under study. Careful analysis of the pedigree and genetic comparison of family members, a typical standard in such genetic susceptibility studies, was not possible due to the absence of family history information in many cases and to the retrospective nature of the study. However, this study is to our knowledge the largest study of patients with MPCs, evaluated using the same multigene targeting panel, and where both germline and multiple tumors DNA were investigated in the highest risk patients.

## Conclusions

eoMPC of colorectal, endometrial, and gastric cancer are considerably enriched for LS patients, supporting the genetic screens of related family members as well as enhanced monitoring for younger patients after their first diagnosis. Routine tumor screening of MSI for patients with initial LS related cancers and consecutive germline testing for patients with MSI-H is worthy to diagnose LS and early detection or avoid second cancer. While susceptibility variants in non-LS genes for MPC patients may exist, evidence for their role is more elusive than for LS patients and would require deeper genetic investigation and complementary functional studies.

## Data Availability

The dataset supporting the conclusions of this article is included within the article and its additional file.

## Abbreviations

MPC: multiple primary cancer
LS: Lynch Syndrome
CPG: cancer predisposition gene
LoF: loss of function
GC: gastric cancer
CRC: colorectal cancer
MSI: microsatellite instability
MMR: mismatch repair
EC: endometrial cancer
eoMPC: early-onset multiple primary cancer
FFPE: formalin fixed paraffin embedded
WES: whole exome sequencing
dMMR: deficiency mismatch repair
P/LP: pathogenic or likely pathogenic
AI: allelic imbalance
LOH: loss of heterozygosity
TCGA: the cancer genome atlas
MAF: minor allele frequency

## Acknowledgements

We would like to thank to Severance Research Initiative project from Yonsei University College of Medicine to support this study.

## Authors’ contributions

YYC, JHC, and OH designed and supervised the research. XX, DN, YYC, JEL generated sequencing data and YYC performed the statistical analysis, prepared the figures and tables, and drafted the manuscript. YYC, JHC, and OH designed the experiments. SS, JEL conducted experiment. SL, JSP, JJ, HK, SHN screened patients and collected specimen. YYC, LM and OH wrote the manuscript. All authors read and approved the final manuscript.

## Ethics approval and consent to participate

This study was approved by Institutional Review Board (IRB) of Severance Hospital and University of California, San Diego (4-2017-0434, 191543). Patient consent was waived as this study is retrospective design.

## Consent for publication

Not applicable

## References

1. Rahman N. Realizing the promise of cancer predisposition genes. Nature. 2014;505:302–8. DOI: 10.1038/nature12981.

2. Huang KL, Mashl RJ, Wu Y, Ritter DI, Wang J, Oh C, et al. Pathogenic Germline Variants in 10,389 Adult Cancers. Cell. 2018;173:355–70 e14. DOI: 10.1016/j.cell.2018.03.039.

3. Evans JP, Powell BC, Berg JS. Finding the Rare Pathogenic Variants in a Human Genome. JAMA. 2017;317:1904–5. DOI: 10.1001/jama.2017.0432.

4. Mersch J, Brown N, Pirzadeh-Miller S, Mundt E, Cox HC, Brown K, et al. Prevalence of Variant Reclassification Following Hereditary Cancer Genetic Testing. JAMA. 2018;320:1266–74. DOI: 10.1001/jama.2018.13152.

5. Cybulski C, Nazarali S, Narod SA. Multiple primary cancers as a guide to heritability. Int J Cancer. 2014;135:1756–63. DOI: 10.1002/ijc.28988.

6. Vogt A, Schmid S, Heinimann K, Frick H, Herrmann C, Cerny T, et al. Multiple primary tumours: challenges and approaches, a review. ESMO Open. 2017;2:e000172. DOI: 10.1136/esmoopen-2017-000172.

7. Whitworth J, Hoffman J, Chapman C, Ong KR, Lalloo F, Evans DG, et al. A clinical and genetic analysis of multiple primary cancer referrals to genetics services. Eur J Hum Genet. 2015;23:581–7. DOI: 10.1038/ejhg.2014.157.

8. Hong S, Won YJ, Park YR, Jung KW, Kong HJ, Lee ES, et al. Cancer Statistics in Korea: Incidence, Mortality, Survival, and Prevalence in 2017. Cancer Res Treat. 2020;52:335–50. DOI: 10.4143/crt.2020.206.

9. Cho I, An JY, Kwon IG, Choi YY, Cheong JH, Hyung WJ, et al. Risk factors for double primary malignancies and their clinical implications in patients with sporadic gastric cancer. Eur J Surg Oncol. 2014; 40:338–44. DOI: 10.1016/j.ejso.2013.10.027.

10. Yun HR, Yi LJ, Cho YK, Park JH, Cho YB, Yun SH, et al. Double primary malignancy in colorectal cancer patients--MSI is the useful marker for predicting double primary tumors. Int J Colorectal Dis. 2009 ;24:369–75. DOI: 10.1007/s00384-008-0541-x.

11. Cancer Genome Atlas Research N. Comprehensive molecular characterization of gastric adenocarcinoma. Nature. 2014;513:202–9. DOI: 10.1038/nature13480.

12. Cancer Genome Atlas N. Comprehensive molecular characterization of human colon and rectal cancer. Nature. 2012;487:330–7. DOI: 10.1038/nature11252.

13. Choi YY, Kim H, Shin SJ, Kim HY, Lee J, Yang HK, et al. Microsatellite Instability and Programmed Cell Death-Ligand 1 Expression in Stage II/III Gastric Cancer: Post Hoc Analysis of the CLASSIC Randomized Controlled study. Ann Surg. 2019;270:309–16. DOI: 10.1097/SLA.0000000000002803.

14. Kim CG, Ahn JB, Jung M, Beom SH, Kim C, Kim JH, et al. Effects of microsatellite instability on recurrence patterns and outcomes in colorectal cancers. Br J Cancer. 2016;115:25–33. DOI: 10.1038/bjc.2016.161.

15. Guastadisegni C, Colafranceschi M, Ottini L, Dogliotti E. Microsatellite instability as a marker of prognosis and response to therapy: a meta-analysis of colorectal cancer survival data. Eur J Cancer. 2010;46 :2788–98. DOI: 10.1016/j.ejca.2010.05.009.

16. Cancer Genome Atlas Research N, Kandoth C, Schultz N, Cherniack AD, Akbani R, Liu Y, et al. Integ rated genomic characterization of endometrial carcinoma. Nature. 2013;497:67–73. DOI: 10.1038/nature12113.

17. Moller P, Seppala T, Bernstein I, Holinski-Feder E, Sala P, Evans DG, et al. Cancer incidence and survival in Lynch syndrome patients receiving colonoscopic and gynaecological surveillance: first report fro m the prospective Lynch syndrome database. Gut. 2017;66:464–72. DOI: 10.1136/gutjnl-2015-309675.

18. Koornstra JJ, Mourits MJ, Sijmons RH, Leliveld AM, Hollema H, Kleibeuker JH. Management of extra colonic tumours in patients with Lynch syndrome. Lancet Oncol. 2009;10:400–8. DOI: 10.1016/S1470-2045(09)70041-5.

19. Cohen SA, Pritchard CC, Jarvik GP. Lynch Syndrome: From Screening to Diagnosis to Treatment in th e Era of Modern Molecular Oncology. Annu Rev Genomics Hum Genet. 2019;20:293–307. DOI: 10.1146/annurev-genom-083118-015406.

20. Pearlman R, Frankel WL, Swanson B, Zhao W, Yilmaz A, Miller K, et al. Prevalence and Spectrum of Germline Cancer Susceptibility Gene Mutations Among Patients With Early-Onset Colorectal Cancer. JAMA Oncol. 2017;3:464–71. DOI: 10.1001/jamaoncol.2016.5194.

21. Umar A, Boland CR, Terdiman JP, Syngal S, de la Chapelle A, Ruschoff J, et al. Revised Bethesda Guidelines for hereditary nonpolyposis colorectal cancer (Lynch syndrome) and microsatellite instability. J Natl Cancer Inst. 2004;96:261–8. DOI: 10.1093/jnci/djh034.

22. Niu B, Ye K, Zhang Q, Lu C, Xie M, McLellan MD, et al. MSIsensor: microsatellite instability detection using paired tumor-normal sequence data. Bioinformatics. 2014;30:1015–6. DOI: 10.1093/bioinformatics/btt755.

23. Eoh KJ, Kim JE, Park HS, Lee ST, Park JS, Han JW, et al. Detection of Germline Mutations in Patients with Epithelial Ovarian Cancer Using Multi-gene Panels: Beyond BRCA1/2. Cancer Res Treat. 2018;50:917–25. DOI: 10.4143/crt.2017.220.

24. Li MM, Datto M, Duncavage EJ, Kulkarni S, Lindeman NI, Roy S, et al. Standards and Guidelines for t he Interpretation and Reporting of Sequence Variants in Cancer: A Joint Consensus Recommendation o f the Association for Molecular Pathology, American Society of Clinical Oncology, and College of American Pathologists. J Mol Diagn. 2017;19:4–23. DOI: 10.1016/j.jmoldx.2016.10.002.

25. https://doi.org/10.5281/zenodo.3743344.D.

26. Van der Auwera GA, Carneiro MO, Hartl C, Poplin R, Del Angel G, Levy-Moonshine A, et al. From FastQ data to high confidence variant calls: the Genome Analysis Toolkit best practices pipeline. Curr Protoc Bioinformatics. 2013;43:1101–033. DOI: 10.1002/0471250953.bi1110s43.

27. DePristo MA, Banks E, Poplin R, Garimella KV, Maguire JR, Hartl C, et al. A framework for variation discovery and genotyping using next-generation DNA sequencing data. Nat Genet. 2011;43:491–8. DOI: 10.1038/ng.806.

28. Wang K, Li M, Hakonarson H. ANNOVAR: functional annotation of genetic variants from high-throughput sequencing data. Nucleic Acids Res. 2010;38:e164. DOI: 10.1093/nar/gkq603.

29. Rentzsch P, Witten D, Cooper GM, Shendure J, Kircher M. CADD: predicting the deleteriousness of variants throughout the human genome. Nucleic Acids Res. 2019;47:D886–D94. DOI: 10.1093/nar/gky1016.

30. Karczewski KJ, Weisburd B, Thomas B, Solomonson M, Ruderfer DM, Kavanagh D, et al. The ExAC browser: displaying reference data information from over 60 000 exomes. Nucleic Acids Res. 2017;45: D840–D5. DOI: 10.1093/nar/gkw971.

31. Landrum MJ, Lee JM, Riley GR, Jang W, Rubinstein WS, Church DM, et al. ClinVar: public archive o f relationships among sequence variation and human phenotype. Nucleic Acids Res. 2014;42:D980–5. DOI: 10.1093/nar/gkt1113.

32. Buckley AR, Ideker T, Carter H, Harismendy O, Schork NJ. Exome-wide analysis of bi-allelic alterations identifies a Lynch phenotype in The Cancer Genome Atlas. Genome Med. 2018;10:69. DOI: 10.1186/s13073-018-0579-5.

33. McLaren W, Gil L, Hunt SE, Riat HS, Ritchie GRS, Thormann A, et al. The Ensembl Variant Effect Predictor. Genome Biology. 2016;17:122. DOI: 10.1186/s13059-016-0974-4.

34. MacArthur DG, Balasubramanian S, Frankish A, Huang N, Morris J, Walter K, et al. A systematic survey of loss-of-function variants in human protein-coding genes. Science. 2012;335:823–8. DOI: 10.1126/science.1215040.

35. Talevich E, Shain AH, Botton T, Bastian BC. CNVkit: Genome-Wide Copy Number Detection and Visualization from Targeted DNA Sequencing. PLoS Comput Biol. 2016;12:e1004873. DOI: 10.1371/journal.pcbi.1004873.

36. Vattathil S, Scheet P. Haplotype-based profiling of subtle allelic imbalance with SNP arrays. Genome R es. 2013;23:152–8. DOI: 10.1101/gr.141374.112.

37. Knudson AG Jr., Mutation and cancer: statistical study of retinoblastoma. Proc Natl Acad Sci U S A. 1971;68:820–3. DOI: 10.1073/pnas.68.4.820.

38. Harismendy O, Kim J, Xu X, Ohno-Machado L. Evaluating and sharing global genetic ancestry in biomedical datasets. J Am Med Inform Assoc. 2019;26:457–61. DOI: 10.1093/jamia/ocy194.

39. Buckley AR, Standish KA, Bhutani K, Ideker T, Lasken RS, Carter H, et al. Pan-cancer analysis reveals technical artifacts in TCGA germline variant calls. BMC Genomics. 2017;18:458. DOI: 10.1186/s12864-017-3770-y.

40. Shin YK, Heo SC, Shin JH, Hong SH, Ku JL, Yoo BC, et al. Germline mutations in MLH1, MSH2 and MSH6 in Korean hereditary non-polyposis colorectal cancer families. Hum Mutat. 2004;24:351. DOI: 10.1002/humu.9277.

41. Desai DC, Lockman JC, Chadwick RB, Gao X, Percesepe A, Evans DG, et al. Recurrent germline muta tion in MSH2 arises frequently de novo. J Med Genet. 2000;37:646–52. DOI: 10.1136/jmg.37.9.646.

42. Karczewski KJ, Francioli LC, Tiao G, Cummings BB, Alfoldi J, Wang Q, et al. The mutational constraint spectrum quantified from variation in 141,456 humans. Nature. 2020;581:434–43. DOI: 10.1038/s41586-020-2308-7.

43. Park S, Supek F, Lehner B. Systematic discovery of germline cancer predisposition genes through the identification of somatic second hits. Nat Commun. 2018;9:2601. DOI: 10.1038/s41467-018-04900-7.

44. Rhees J, Arnold M, Boland CR. Inversion of exons 1-7 of the MSH2 gene is a frequent cause of unexplained Lynch syndrome in one local population. Fam Cancer. 2014;13:219–25. DOI: 10.1007/s10689-013-9688-x.

45. Win AK, Lindor NM, Young JP, Macrae FA, Young GP, Williamson E, et al. Risks of primary extracolonic cancers following colorectal cancer in lynch syndrome. J Natl Cancer Inst. 2012;104:1363–72. DOI: 10.1093/jnci/djs351.

46. Gupta S, Provenzale D, Regenbogen SE, Hampel H, Slavin TP, Hall MJ, et al. NCCN Guidelines Insig hts: Genetic/Familial High-Risk Assessment: Colorectal, Version 3.2017. J Natl Compr Canc Netw. 2017;15:1465–75. DOI: 10.6004/jnccn.2017.0176.

47. Hampel H, Frankel W, Panescu J, Lockman J, Sotamaa K, Fix D, et al. Screening for Lynch syndrome (hereditary nonpolyposis colorectal cancer) among endometrial cancer patients. Cancer Res. 2006;66:7810–7. DOI: 10.1158/0008-5472.CAN-06-1114.

48. Le DT, Uram JN, Wang H, Bartlett BR, Kemberling H, Eyring AD, et al. PD-1 Blockade in Tumors wi th Mismatch-Repair Deficiency. N Engl J Med. 2015;372:2509–20. DOI: 10.1056/NEJMoa1500596.

49. Latham A, Srinivasan P, Kemel Y, Shia J, Bandlamudi C, Mandelker D, et al. Microsatellite Instability Is Associated With the Presence of Lynch Syndrome Pan-Cancer. J Clin Oncol. 2019;37:286–95. DOI: 10.1200/JCO.18.00283.

50. Galvan A, Ioannidis JP, Dragani TA. Beyond genome-wide association studies: genetic heterogeneity a nd individual predisposition to cancer. Trends Genet. 2010;26:132–41. DOI: 10.1016/j.tig.2009.12.008.

51. Pirinen M, Donnelly P, Spencer CC. Including known covariates can reduce power to detect genetic eff ects in case-control studies. Nat Genet. 2012;44:848–51. DOI: 10.1038/ng.2346.

52. Hindorff LA, Sethupathy P, Junkins HA, Ramos EM, Mehta JP, Collins FS, et al. Potential etiologic an d functional implications of genome-wide association loci for human diseases and traits. Proc Natl Aca d Sci U S A. 2009;106:9362–7. DOI: 10.1073/pnas.0903103106.

53. Chen S, Wang W, Lee S, Nafa K, Lee J, Romans K, et al. Prediction of germline mutations and cancer risk in the Lynch syndrome. JAMA. 2006;296:1479–87. DOI: 10.1001/jama.296.12.1479.

54. He YJ, Qiao ZY, Gao B, Zhang XH, Wen YY. Association between RECQL5 genetic polymorphisms a nd susceptibility to breast cancer. Tumour Biol. 2014;35:12201–4. DOI: 10.1007/s13277-014-2528-2.

55. Fewings E, Larionov A, Redman J, Goldgraben MA, Scarth J, Richardson S, et al. Germline pathogenic variants in PALB2 and other cancer-predisposing genes in families with hereditary diffuse gastric cancer without CDH1 mutation: a whole-exome sequencing study. Lancet Gastroenterol Hepatol. 2018;3:489–98. DOI: 10.1016/S2468-1253(18)30079-7.

56. Lin Y, Chen H, Wang X, Xiang J, Wang H, Peng J. Mining the role of RECQL5 in gastric cancer and seeking potential regulatory network by bioinformatics analysis. Exp Mol Pathol. 2020;115:104477. DOI: 10.1016/j.yexmp.2020.104477.

57. Sun T, Gao Y, Tan W, Ma S, Shi Y, Yao J, et al. A six-nucleotide insertion-deletion polymorphism in t he CASP8 promoter is associated with susceptibility to multiple cancers. Nat Genet. 2007;39:605–13. DOI: 10.1038/ng2030.

58. Zhang Y, Li W, Hong Y, Wu G, He K, Liu D. A systematic analysis of the association studies between CASP8 D302H polymorphisms and breast cancer risk. J Genet. 2017;96:283–9. DOI: 10.1007/s12041-017-0774-y.

59. Ying Y, Xu J, Qi Y, Zhang M, Yang Y. CASP8 rs3834129 (−652 6N insertion/deletion) Polymorphism and Colorectal Cancer Susceptibility: An Updated Meta-Analysis. J Cancer. 2018;9:4166–71. DOI: 10.7150/jca.27110.

60. Chen H, Zhang S, Wu Z. Fanconi anemia pathway defects in inherited and sporadic cancers. Transl Pediatr. 2014;3:300–4. DOI: 10.3978/j.issn.2224-4336.2014.07.05.

61. Esteban-Jurado C, Franch-Exposito S, Munoz J, Ocana T, Carballal S, Lopez-Ceron M, et al. The Fanc oni anemia DNA damage repair pathway in the spotlight for germline predisposition to colorectal cancer. Eur J Hum Genet. 2016;24:1501–5. DOI: 10.1038/ejhg.2016.44.

62. Huang JP, Lin J, Tzen CY, Huang WY, Tsai CC, Chen CJ, et al. FANCA D1359Y mutation in a patient with gastric polyposis and cancer susceptibility: A case report and review of literature. World J Gastroenterol. 2018;24:4412–8. DOI: 10.3748/wjg.v24.i38.4412.

63. Soung YH, Lee JW, Kim SY, Jang J, Park YG, Park WS, et al. CASPASE-8 gene is inactivated by som atic mutations in gastric carcinomas. Cancer Res. 2005;65:815–21.

64. Soung YH, Lee JW, Kim SY, Sung YJ, Park WS, Nam SW, et al. Caspase-8 gene is frequently inactivated by the frameshift somatic mutation 1225_1226delTG in hepatocellular carcinomas. Oncogene. 2005;24:141–7. DOI: 10.1038/sj.onc.1208244.

65. Ramanagoudr-Bhojappa R, Carrington B, Ramaswami M, Bishop K, Robbins GM, Jones M, et al. Multiplexed CRISPR/Cas9-mediated knockout of 19 Fanconi anemia pathway genes in zebrafish revealed their roles in growth, sexual development and fertility. PLoS Genet. 2018;14:e1007821. DOI: 10.1371/journal.pgen.1007821.

66. Pulliam-Leath AC, Ciccone SL, Nalepa G, Li X, Si Y, Miravalle L, et al. Genetic disruption of both Fa ncc and Fancg in mice recapitulates the hematopoietic manifestations of Fanconi anemia. Blood. 2010; 116:2915–20. DOI: 10.1182/blood-2009-08-240747.

67. Hashibe M, Boffetta P, Zaridze D, Shangina O, Szeszenia-Dabrowska N, Mates D, et al. Contribution o f tobacco and alcohol to the high rates of squamous cell carcinoma of the supraglottis and glottis in Cen tral Europe. Am J Epidemiol. 2007;165:814–20. DOI: 10.1093/aje/kwk066.

68. Gandini S, Botteri E, Iodice S, Boniol M, Lowenfels AB, Maisonneuve P, et al. Tobacco smoking and cancer: a meta-analysis. Int J Cancer. 2008;122:155–64. DOI: 10.1002/ijc.23033.

69. Services USDoHaH. 14th report on carcinogens. https://ntp.niehs.nih.gov/whatwestudy/assessments/cancer/roc/index.html. 2016.

70. Li X, Yao X, Wang Y, Hu F, Wang F, Jiang L, et al. MLH1 promoter methylation frequency in colorectal cancer patients and related clinicopathological and molecular features. PLoS One. 2013;8:e59064. DOI: 10.1371/journal.pone.0059064.

